# The relationship between schizophrenia polygenic scores, blood-based proteins and psychosis diagnosis in the UK Biobank

**DOI:** 10.1101/2025.08.17.25333857

**Authors:** Kimberley M Kendall, Sophie E Legge, Eilidh Fenner, Peter Holmans, James TR Walters

## Abstract

Despite notable progress in psychiatric genomics, there are no validated blood-based biomarkers for psychosis. Previous studies have failed to establish a link between schizophrenia polygenic scores (PGS) and blood protein levels. We aimed to identify associations between schizophrenia PGS and blood-based proteins, and to determine whether levels of these proteins differ in individuals with psychosis. We analysed proteomic and genomic data from 49,083 participants in the UK Biobank. Association analyses, excluding individuals with psychosis, identified nominal associations (p < 0.05) of schizophrenia PGS with 109 proteins. Four of these (TMPRSS15, ADGRB3, CEACAM21, and KLK1) met the false discovery rate (FDR) threshold of < 0.05. We investigated the association of these four proteins with psychosis in a matched case-control sample (291 cases, 873 controls). In individuals with psychosis, we observed significantly higher levels of TMPRSS15 (effect size 0.22, standard error 0.07, FDR 6.97 × 10^−3^) and lower levels of KLK1 (effect size −0.23, SE 0.09, FDR 3.34 × 10^−2^). These two proteins should be taken forward for further study and validation aimed at investigating their potential as psychosis biomarkers.

## Introduction

Psychosis is an umbrella term for a group of psychiatric disorders characterised by alterations in perceptions, experiences, thoughts and behaviour. Psychotic disorders, such as schizophrenia and schizoaffective disorder, are highly heritable (1), with complex genetic architecture. For example, genetic risk for schizophrenia is conferred by rare variants of large effect, and common variants of individually smaller effect, known as single-nucleotide polymorphisms (SNPs), with associations identified at 287 common variant loci.(2)

Despite considerable advances in psychiatric genomics, there are currently no validated blood-based biomarkers for psychosis. In contrast, biomarker discovery in Alzheimer’s disease has brought considerable benefits, enabling improved diagnosis, prognosis prediction, patient stratification, and therapeutic development.(3–5) Numerous studies have reported alterations in blood-based proteins in psychotic disorders, particularly those involved in immune and inflammatory processes.(6–11) However, it is unclear whether these changes reflect genetic liability, disease pathology or treatment effects. The limited research investigating associations between schizophrenia genetic liability, indexed by schizophrenia polygenic scores, and blood-based proteins reported no association.(6)

In this study, we aimed to identify blood-based biomarkers for psychosis by investigating proteins associated with schizophrenia PGS. We then tested whether levels of these proteins differed in individuals with psychosis compared to controls.

## Methods

### Participants

Individuals aged 40–69 years living in the United Kingdom (UK) were recruited to the UK Biobank between 2006 and 2010. Participants attended UK Biobank assessment centres where they provided detailed health and lifestyle information, underwent cognitive assessments, and supplied biological samples, including blood.(12) The North West Multi-Centre Ethics Committee granted ethical approval to the UK Biobank, and all participants gave written informed consent. This study was conducted under the UK Biobank project number 13310.

### Genotyping

The UK Biobank participant samples were genotyped at the Affymetrix Research Services Laboratory, Santa Clara, CA. Genotyping was conducted on two arrays, between which there was 95% common content: the UK Biobank Axiom array and the UK BiLEVE array. Standard sample processing was applied and is described elsewhere (13,14), before imputation using the Haplotype Reference Consortium Panel.(15) Post-imputation QC included a minor allele frequency greater than 0.01, an imputation score greater than 0.8, missingness of less than 0.05, and a Hardy-Weinberg equilibrium *P* value greater than 1 × 10^−6^.(16,17)

### Genetic Ancestry

Genetic ancestries were inferred using the UKBB array data. The process is described in detail elsewhere (18) and was based on previously published methods.(19,20) In summary, principal components were computed in the 1000 Genomes Phase 3 reference based on 65,338 ancestry-informative markers. The samples were projected into reference PC space, and the top 34 PCs were input into a Fisher’s Linear Discriminant Analysis model to assign genetic ancestry probabilities. Individuals were assigned a 1KGP-like genetic ancestry label if their ancestry probability exceeded 0.8.(18) We note that these labels reflect genetic similarity to the 1KGP reference panel and do not represent discrete or biologically distinct categories. All analyses in this paper were conducted on participants with a 1KGP-European-like ancestry probability of 0.8 or higher. The first five principal components from the projection were used as covariates to control for population stratification.

### Polygenic Scores

Polygenic scores (PGS) for schizophrenia were calculated based on genome-wide summary statistics from a custom GWAS of schizophrenia from the Psychiatric Genomics Consortium (PGC) (2) that excluded UK Biobank participants.(21) The PGS were derived using a clumping and thresholding approach, consistent with the PGC (2), in PRSice-2.(22) Only SNPs with a p-value of < 0.05 were included. Further details are described elsewhere.(21)

### Proteomic Measurement, Processing and Quality Control

The UK Biobank collected blood samples from participants during their attendance at assessment centres between 2006 and 2010. A total of 54,219 UK Biobank participants were selected for proteomic analysis as part of the UK Biobank Pharma Proteomics Project, including i) 46,595 randomly selected participants who provided blood samples at their initial baseline visit, ii) 1,268 participants from a COVID-19 repeat imaging study, and 6,376 participants selected due to specific characteristics, such as case status for coronary heart disease, diabetes mellitus, and chronic kidney disease (groups are not mutually exclusive).(23)

The UK Biobank processed samples centrally to extract plasma, which was then stored at −80°C before analysis. Proteomic analyses were conducted on the Olink Explore 3072 platform and are reported in normalised protein expression (NPX) units. NPX is an arbitrary unit on a log2 scale – a one-unit difference corresponds to a doubling of protein concentration. These values are relative and internally normalised across samples and plates to allow comparison of protein levels. Samples were processed in batches, with quality control procedures used to mitigate batch effects, including median signal normalisation within each plate.(23) The UK Biobank provided proteomic data for 53,013 participants. Additionally, we excluded i) participants with more than 60% missing protein values, ii) proteins with more than 60% missing data, and iii) proteins with more than 20% of values below the limit of detection. This resulted in a final set of 2,077 proteins for analysis (Supplementary Table 1).

### Phenotypic Definitions

We defined psychosis diagnosis based on the first occurrence of the diagnosis, as collated by the UK Biobank from death registers, primary care records, hospital admission data, and self-reports (field 130875, accessed March 2025). This included diagnoses of schizophrenia (International Classification of Diseases 10, ICD-10, code F20), persistent delusional disorder (F22), schizoaffective disorder (F25) and unspecified nonorganic psychosis (F29).(24)

We extracted a list of antipsychotic medications from the UK Biobank data (field 20003). We used these to create a binary variable indicating whether participants were prescribed antipsychotic medication at the time of their attendance at the UK Biobank assessment centres (Supplementary Table 2).

### Statistical Analyses

All analyses were conducted on the UK Biobank Research Analysis Platform using R (version 4.4.0). In the primary analysis, we examined associations between schizophrenia PGS and the levels of 2,077 proteins. Covariates included sex (field 31), age (field 21003), age squared, time between sample collection (field 53) and analysis (UKBB proteomic files), proteomic analysis batch (UKBB proteomic files), the first five genetic principal components, BMI (field 21001), current smoking status (field 20116), medication (number of medications, field 137), renal function (eGFR calculated using fields 30700, 30720 in addition to age and sex), and liver function (ALT, field 30620). Individuals with diagnoses of psychosis were excluded from these analyses. To account for multiple testing, we adjusted p-values using the Benjamini-Hochberg false discovery rate (FDR) correction applied across all 2,077 proteins, with a significance threshold of 0.05.

To further investigate the four PGS-associated proteins that met the FDR of < 0.05 threshold in the primary analysis, we constructed a matched case-control cohort of individuals with and without diagnoses of psychosis. Matching was conducted using the MatchIt package (25) in a 3:1 ratio, based on age, sex, ethnicity, interval between sample acquisition and analysis, current smoking status, BMI and number of medications (291 cases, 873 controls). We then conducted linear regression analyses with protein level as the dependent variable and psychosis diagnosis as the predictor. We, once again, accounted for multiple testing using the Benjamini-Hochberg FDR method across the five proteins, with a significance threshold of 0.05.

We then conducted follow-up analyses to determine whether antipsychotic medication prescription was associated with the levels of the two proteins identified as differing between individuals with diagnoses of psychosis and those without. To achieve this, we performed linear regression analyses with antipsychotic medication as the predictor, protein level as the outcome, and age, sex, and schizophrenia PGS as covariates.

## Results

### Association analyses of schizophrenia PGS and blood-based protein levels

Following quality control procedures, ancestry restriction and the exclusion of 291 individuals with diagnoses of psychosis, we retained data for 48,792 individuals across 2,077 proteins. 54% of the participants were female (n = 26,361), and the mean age of the participants at the time of sample collection (UKBB first assessment centre visit) was 57.1 years (range 40 – 70 years).

We performed linear regression analyses to test for associations between schizophrenia PGS and protein levels. Of the 2,077 proteins analysed, 109 were associated with schizophrenia PGS at nominal levels of significance (p < 0.05, Supplementary Table 1) and four met the FDR < 0.05 threshold (Table 1).

**Table 1.** Association of schizophrenia PGS with blood protein levels in individuals without diagnoses of psychosis. Results are shown for associations achieving significance at an FDR < 0.05. SE – standard error; FDR – false discovery rate.

| Protein | Effect Size (SE) | FDR |
| --- | --- | --- |
| TMPRSS15<br>Transmembrane Serine Protease 15 | -149.5 (33.8) | $6.73 \times 10^{-3}$ |
| ADGRB3<br>Adhesion G Protein-Coupled Receptor B3 | 63.4 (12.5) | $3.73 \times 10^{-4}$ |
| CEACAM21<br>CEA Cell Adhesion Molecule 21 | 135.9 (31.9) | $1.04 \times 10^{-2}$ |
| KLK1<br>Kallikrein 1 | 330.5 (47.8) | $1.00 \times 10^{-8}$ |

**Table 2.** Association of blood-based protein levels with psychosis diagnosis in a matched case-control cohort. SE – standard error; FDR – false discovery rate.

| Protein | Effect Size (SE) | FDR |
| --- | --- | --- |
| TMPRSS15<br>Transmembrane Serine Protease 15 | 0.17 (0.07) | $3.34 \times 10^{-2}$ |
| ADGRB3<br>Adhesion G Protein-Coupled Receptor B3 | 0.02 (0.03) | 0.47 |
| CEACAM21<br>CEA Cell Adhesion Molecule 21 | -0.10 (0.07) | 0.19 |
| KLK1<br>Kallikrein 1 | -0.23 (0.09) | $3.34 \times 10^{-2}$ |

### Examining differences in protein levels in individuals with diagnoses of psychosis

A total of 0.59% (n = 291) of our original 49,083 participants had a diagnosis of psychosis. We examined the association between psychosis diagnosis and levels of the four proteins that met the FDR threshold in the primary analysis to determine if their levels differed in individuals with diagnoses of psychosis compared to controls. We conducted this analysis in a matched psychosis diagnosis case-control cohort (291 cases, 873 controls). Two proteins were present at significantly different levels in individuals with diagnoses of psychosis: i) TMPRSS15 was present at elevated levels (effect size 0.17, SE 0.07, FDR 3.34 × 10^−2^), and ii) KLK1 was present at lower levels (effect size −0.23, SE 0.09, FDR 3.34 × 10^−2^), in individuals with diagnoses of psychosis.

The directions of effect for both TMPRSS15 and KLK1 differed between the association analyses of schizophrenia PGS with protein levels and the analysis of associations between protein levels and psychosis diagnosis. We investigated whether this pattern of results may have occurred due to the effect of antipsychotic medication. Medication data were available on 78.15% of the cohort (n = 38,358). Of these individuals, 434 were treated with antipsychotic medication (Supplementary Table 2), and 29.49% (n = 128) of these had a diagnosis of psychosis. We conducted a linear regression analysis with schizophrenia PGS, psychosis diagnosis and antipsychotic medication prescription as predictors; age, sex, batch and time delay as covariates; and protein levels as outcomes. For TMPRSS15 and KLK1, both schizophrenia PGS and psychosis diagnosis were associated with protein levels, but antipsychotic prescription was not (Supplementary Table 3).

## Discussion

In this study, we explored the relationship between schizophrenia PGS and blood-based protein levels in the UK Biobank, a large, population-based cohort. Of the 2,077 blood-based proteins analysed, we identified 109 whose blood levels were nominally associated with schizophrenia PGS (Supplementary Table 1), four of which met the FDR threshold in analyses adjusted for multiple potential confounders - TMPRSS15, transmembrane serine protease 15; ADGRB3, adhesion G protein-coupled receptor B3; CEACAM21, CEA Cell Adhesion Molecule 21; and KLK1, kallikrein 1.

We then examined whether the candidate proteins showed different levels in individuals with diagnoses of psychosis by analysing a psychosis diagnosis case-control sub-cohort from the UK Biobank. Two proteins, TMPRSS15 and KLK1, exhibited altered levels in individuals with psychosis compared to those without: TMPRSS15 levels were elevated, while KLK1 levels were decreased.

Transmembrane serine protease 15 (TMPRSS15), also known as enteropeptidase, belongs to the serine protease family, and its role is to activate trypsinogen to trypsin during digestion.(26) Although primarily expressed in the small intestine, there is evidence of low-level expression in the brain, particularly in neurons under stress conditions. In 2010, Makarova et al demonstrated in a rat model that TMPRSS15 is expressed in hippocampal neurons, where it modulates neuronal survival under glutamate toxicity.(27) Other members of the serine protease family play roles in synaptic plasticity and neuroinflammation.(28,29) SNPs within the *TMPRSS15* gene have previously been reported in GWAS as associated with caudal anterior cingulate cortex volume (psychosis neuroimaging study)(30), with nucleus accumbens volume in trauma-exposed individuals (31), and with educational attainment.(32) Examination of the summary statistics from the largest schizophrenia GWAS to date revealed 35 SNPs within the *TMPRSS15* gene associated with schizophrenia at nominal levels of significance (lowest p value 1.23 × 10^−3^). For these associations, 28 of the effect directions were negative and seven were positive.(2)

Kallikrein-1 (KLK1) is a member of the kallikrein family of serine proteases. It cleaves low-molecular-weight kininogen to produce bradykinin, a vasoactive peptide which plays roles in vasodilatation, inflammation modulation, neovascularisation and protection against ischaemic injury in cardiovascular and renal tissue.(33) KLK1 is expressed in a range of tissues, including vasculature, neutrophils and the central nervous system (cerebral cortex and hippocampus).(34) To our knowledge, KLK1 has not been implicated previously in schizophrenia, and a review of the schizophrenia GWAS summary statistics revealed only a suggestive association with the rs2659058 SNP (Beta 0.01509549, SE 0.0081, p 0.06324).(2) KLK1 has been reported to be neuroprotective in cases of cerebral ischaemia (33), to decrease depression-like behaviours in rodent models (35) and to be dysregulated in multiple sclerosis.(36) In addition, the broader family of kallikreins have been implicated in schizophrenia, bipolar disorder and depression, as well as several neurological disorders, including Parkinson’s disease, Alzheimer’s disease, multiple sclerosis and epilepsy.(37)

In our study, for both TMPRSS15 and KLK1, the direction of the association was opposite when comparing the primary analysis (association of schizophrenia PGS with protein levels) and the follow-up analyses (association of the protein levels with psychosis diagnosis). Schizophrenia PGS was associated with lower levels of TMPRSS15, but TMPRSS15 levels were higher in individuals with diagnoses of psychosis. Schizophrenia PGS was associated with higher levels of KLK1, but KLK1 levels were lower in individuals with psychosis. The potential explanations for this pattern of results include i) the proteins may be up- or down-regulated in response to the disease process itself, and ii) the proteins may have altered levels in individuals with diagnoses of psychosis due to the effects of medication. We examined this latter explanation by conducting association analyses of antipsychotic medication, schizophrenia PGS and psychosis diagnosis simultaneously with protein levels. For both proteins, schizophrenia PGS and psychosis diagnosis remained associated with the levels of TMPRSS15 and KLK1, but antipsychotic medication prescription was not associated (Supplementary Table 3).

We used schizophrenia PGS to investigate associations with blood-based proteins in individuals without diagnoses of psychosis. This method is potentially more directly linked to inherited susceptibility than restricting analyses to case-control samples, where outcomes may be influenced by illness status or treatment effects. Earlier studies, including those using similar proteomic platforms (e.g., Olink), did not find strong connections between polygenic scores and protein levels, possibly due to limited statistical power. Our use of a large, well-characterised cohort allowed us to detect small but statistically significant effects.

Further research should explore the potential of TMPRSS15 and KLK1 as biomarkers for psychosis. Ideally, this would involve individuals at high risk of psychosis due to genetic factors or those diagnosed with the At Risk Mental State to evaluate the proteins’ diagnostic and predictive value, as well as individuals experiencing their first episode of psychosis with longitudinal follow-up data to determine prognostic utility.

Our results should be interpreted in light of several limitations. First, our observed effect sizes were small. Second, the number of individuals with diagnoses of psychosis in the UK Biobank is relatively low, which may limit power for downstream case-control analyses. In addition, individuals with diagnoses of psychosis in this cohort are unusually well functioning, and this may limit the generalisability of the results.(21) Finally, protein levels were measured cross-sectionally, and in most cases, after a diagnosis of psychosis was made. Our findings should ideally be confirmed through longitudinal or experimental studies, with data collected in a manner that allows for the temporal relationship between protein level alterations and diagnosis to be established, thereby permitting causal analyses.

In conclusion, we identified multiple blood-based proteins associated with schizophrenia PGS. We provide preliminary evidence that two of these proteins, TMPRSS15 and KLK1, have significantly altered levels in individuals with diagnoses of psychosis, although the direction of effect was opposite to that observed in the primary analysis. These results were not explained by antipsychotic medication prescription. These findings highlight new avenues for understanding the biological pathways connecting genetic liability to psychosis diagnosis and potential biomarkers for future research.

## Data Availability

All data used in the present study are available from the UK Biobank.

